# COVID-19 is an emergent disease of aging

**DOI:** 10.1101/2020.04.15.20060095

**Authors:** Didac Santesmasses, José Pedro Castro, Aleksandr A. Zenin, Anastasia V. Shindyapina, Maxim V. Gerashchenko, Bohan Zhang, Csaba Kerepesi, Sun Hee Yim, Peter O. Fedichev, Vadim N. Gladyshev

## Abstract

COVID-19 is an ongoing pandemic caused by the SARS-CoV-2 coronavirus that poses one of the greatest challenges to public health in recent years. SARS-CoV-2 is highly contagious and often leads to severe viral pneumonia with respiratory failure and death in the elderly and subjects with pre-existing conditions, but the reason for this age dependence is unclear. Here, we found that the case fatality rate for COVID-19 grows exponentially with age in Italy, Spain, South Korea, and China, with the doubling time approaching that of all-cause human mortality. In addition, men and those with multiple age-related diseases are characterized by increased mortality. Moreover, similar mortality patterns were found for all-cause pneumonia. We further report that the gene expression of ACE2, the SARS-CoV-2 receptor, grows in the lung with age, except for subjects on a ventilator. Together, our findings establish COVID-19 as an emergent disease of aging, and age and age-related diseases as its major risk factors. In turn, this suggests that COVID-19, and deadly respiratory diseases in general, may be targeted, in addition to therapeutic approaches that affect specific pathways, by approaches that target the aging process.

## Introduction

Our society faces unprecedented challenges as the global pandemic of the Coronavirus Disease 2019 (COVID-19) spreads around the world, with more than one million people infected and almost hundred thousand confirmed deaths^1^. The disease is caused by an enveloped single-stranded positive RNA virus named severe acute respiratory syndrome coronavirus 2 or SARS-CoV-2^2,3^. In contrast to other coronaviruses, SARS-CoV-2 has the ability to infiltrate the lower respiratory tract resulting in severe lung damage and a high rate of deaths from pneumonia ^4^.

A particular characteristic of SARS-CoV-2 is its higher prevalence among older people and those with pre-existing conditions such as hypertension, diabetes, cancer, heart failure and chronic obstructive pulmonary disease. These individuals show a much higher susceptibility to the disease and poor clinical prognosis, and as a result, lower chance of survival. Interestingly, the comorbidities have aging as a common factor and have been described in recent years as age-related diseases. A recent study reported estimates of the age-stratified case fatality rate and showed it to be lower in those below 60 years old (1,4% [0,4–3,5]) compared to subjects who were 60 years or older (4,5% [1,8–11,1])^5^. However, the reason the elderly and those with pre-existing conditions display a higher risk for COVID-19 is currently unknown.

In the lung, where SARS-CoV-2 docks and initiates infection, this susceptibility might be related to changes in the physical properties of the tissue and/or aging of the immune function known as immunosenescence. The lung employs mechanical defenses such as cough, the barrier function of the mucus and epithelium, and mucociliary clearance, which in sync with the innate immune system, help to clear aspirated or inhaled substances, including infectious agents^6^. However, these concerted actions that avoid the deposit and infection of harmful substances and pathogens are known to decrease with aging.

In general, the idea that older people are more susceptible to infections is not new. In fact, it has been reported that up to one third of deaths in the elderly is a result of infectious diseases^7^. Persistent viral infections may also trigger monoclonal expansion of T cells, which over the lifetime results in poor variability of memory T-cells. In turn, this eventually drives immune exhaustion due to the decline in T cell diversity^8^, a critical problem when facing novel threats such as SARS-CoV-2.

An additional feature that characterizes the severe cases of COVID-19 is the elevated levels of inflammation that can compromise lung tissue integrity and function, leading to pneumonia. Remarkably, accumulated and exhausted T cells secrete preferentially pro-inflammatory cytokines such as IFN and TNF. These cytokines can contribute, along with the innate immune system, to the low grade pro-inflammatory background observed in the elderly^9^, which may worsen COVID-19 outcomes and explain the elevated levels of inflammation. It is also possible that age-associated clonal hematopoiesis may contribute to the increased inflammation due to hematopoietic stem cell myeloid generation bias of pro-inflammatory macrophages and mast cells, and reduction of lymphoid differentiation^10^. Moreover, decreased T-cell capacity to properly activate antibody-secreting cells to further elicit effective immune responses may be compromised^11^. Yet, another possible explanation is thymus involution^12^. During aging, the thymus becomes atrophic and is gradually replaced by fibrotic tissue^13^. This results in a reduced number, or even complete abrogation, of exiting naive T cells^14^. In support of the importance of thymus involution, recent studies show that the age-stratified infection rate of COVID-19 correlates with involution of the thymus^15^. Together, all these features may result in the decreased ability of older people to fight viral infections, leading to age-related inflammation and higher susceptibility of the lung to the COVID-19-inflicted damage.

From a molecular perspective, SARS-CoV-2 selectivity towards the elderly may be explained by alterations in the transcriptome, proteome or metabolome, or in certain key genes, proteins or metabolites. SARS-CoV-2 has been found to infect host cells by attaching its viral spike (S) protein to a receptor named angiotensin-converting enzyme 2 (ACE2). This binding seems to be highly specific and facilitates the virus entry via endocytosis^16^. Therefore, the level of ACE2 expression may be a factor in COVID-19 infection. Expression of this protein is known to be affected by diverse stimuli and certain drugs used to treat hypertension, diabetes, and other diseases. However, how ACE2 expression is regulated by aging, and how it is affected in the lung, the site of SARS-CoV-2 infection, is not known.

In this work, we revealed a strong link between COVID-19 and aging. Based on our analysis, we propose that COVID-19, and more generally deadly respiratory diseases, should be considered as novel and emergent diseases of aging. Understanding that age is a major factor for COVID-19 may help to design approaches against this disease that target the aging process, along with specific antiviral approaches and those that boost more efficiently the human immune system of the elderly and those with comorbidities.

## Results

We analyzed the data on SARS-CoV-2 infections for Italy, where most of the fatalities occurred as of March 26, 2020^17^. In total, 73,780 cases were reported positive for SARS-CoV-2 with the median age of patients being 62 years. 88% of the infected individuals were 40 years or older (Fig. 1A); however, there was no consistent trend towards an increased incidence rate in older people. 6,801 patients had reportedly died, with 95% of the fatalities observed in subjects 60 years old or older (Fig. 1B). More men than women were reported to be infected (57.6% and 42.4%, respectively) and to have died from COVID-19 (70.4% and 29.6%, respectively). The gender difference for infections and mortality was consistent across all age groups except for the group of 90 years or older, where confirmed cases and fatalities were higher in women, which may be explained by the population structure (i.e., more women survive to old age).

**Fig. 1.**
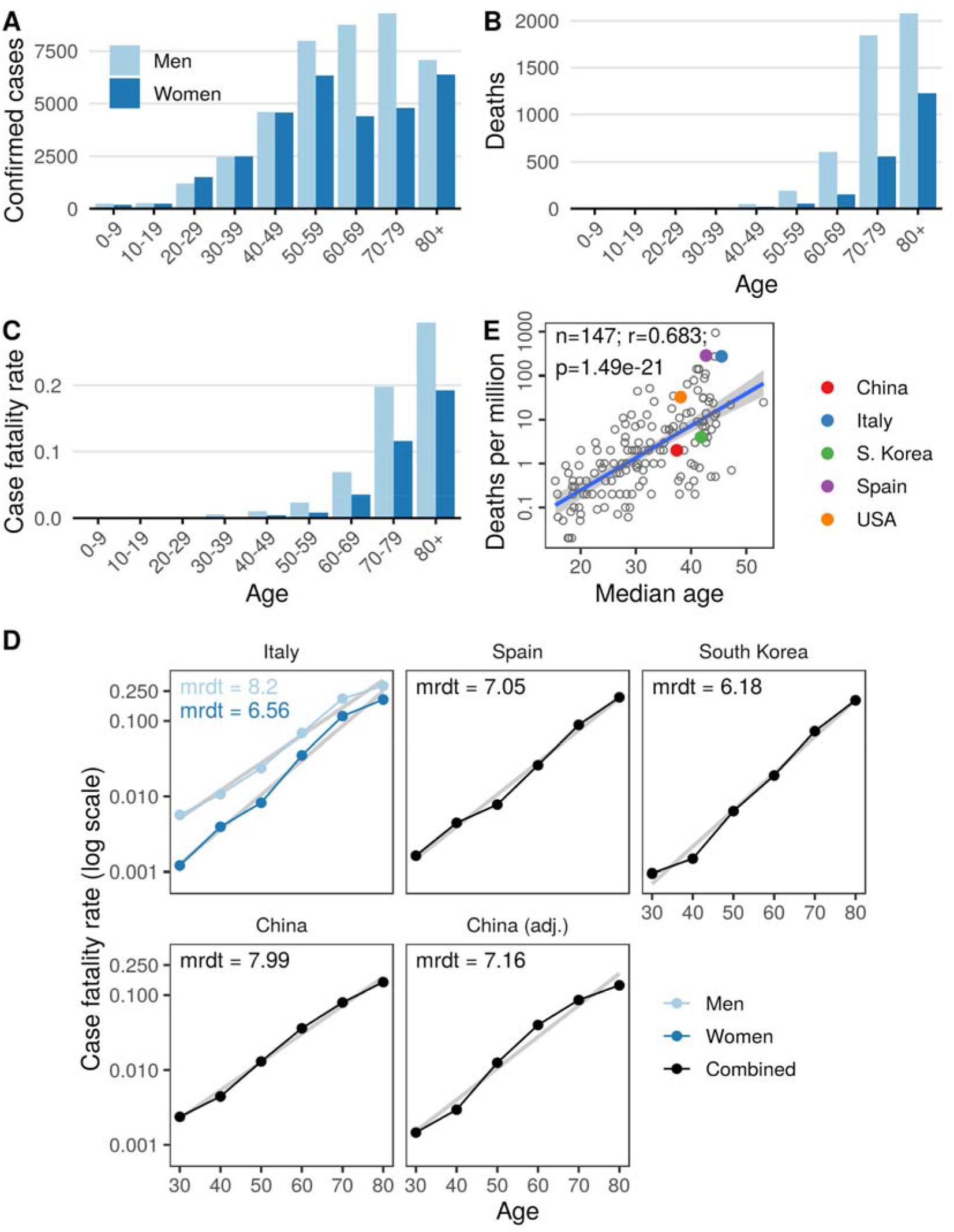
Age is a major risk factor for COVID-19. (A) Confirmed cases of COVID-19 in Italy. Data are through March 26, 2020. (B) Deaths in Italy. Men are in light blue, and women in dark blue. (C) Case fatality rate in men and women across age groups. (D) Mortality rate exponentially increases with age. Case fatality rate as a function for indicated age groups and countries. Linear regression of log of case fatality rate on age is shown by light lines. Men are shown in light blue and women in dark blue. Combined cases (men + women) are shown in black for indicated countries. mrdt - mortality rate doubling time. (E) COVID-19 mortality as a function of the median age of the country. The countries discussed in the study are labeled.

The case fatality rate (CFR), i.e. the quotient of deaths to confirmed infections, is commonly used to assess disease severity, and to estimate the outcome of the disease. The CFR of COVID-19 exponentially grows with age in both genders, exceeding 20% in people above 80 years old (Fig. 1C), whereas it is low in young adults and children (Supplementary Fig. 1 and Supplementary Fig. 2). This pattern is reminiscent of all-cause mortality in the human population, which doubles approximately every 8 years, also known as mortality rate doubling time (MRDT). Based on the COVID-19 CFR data, we assessed the doubling time of COVID-19 deaths using the Gompertz law (Fig. 1D). The doubling time was 8.2 years for men, and 6.6 years for women. We then extended the analyses to other countries and found that the doubling times ranged from 6.18 to 7.99 years for Spain and China, two other countries heavily affected by COVID-19, and for South Korea, which carried out an extensive testing program (Fig. 1D).

Estimation of CFR of an infectious disease during an outbreak is challenging. First, there is a time lapse between a person developing the symptoms, the case being detected and reported, and the outcome of the disease. Second, the detection of a newly emerged pathogen is biased towards clinically severe cases. Therefore, naive CFR (ratio of fatalities over confirmed cases) can be somewhat inaccurate in the assessment of disease severity, which could explain differences observed between different countries. Estimates of CFR attempting to correct for these effects have been produced by several groups^5,18,19^. Here, we used the CFR estimates adjusting for underlying demography and potential under-ascertainment of mainland China^5^. This allowed us to compare the doubling time of naive CFR and adjusted CFR. The doubling time obtained using the adjusted CFR was 7.16 years, approximately 10% shorter than the naive CFR. Nonetheless, the doubling time using adjusted CFR was not substantially different compared to other countries (Fig. 1D). We further found that the rate of death from COVID-19 (adjusted to population size) grows exponentially with the median age of the country (Fig. 1E), presumably because countries with older populations have a higher fraction of people who may succumb to this disease. The incidence of COVID-19 also grows exponentially with the median age of the country (Supplementary Fig. 3). Overall, the data show that not only CFR for COVID-19 grows exponentially with age, but its doubling time approaches that of all-cause human mortality. Together with a higher mortality for men than for women, this finding establishes COVID-19 as a disease of aging, with age emerging as a major risk factor for COVID-19.

The vast majority of deaths from COVID-19 were observed in patients with pre-existing conditions. Based on a sample of 710 deceased patients with detailed medical history, only 2.1% of deaths occurred with no comorbidities diagnosed. The most common comorbidity was hypertension, diagnosed in 70% of the patients, followed by diabetes (31%) (Fig. 2A). The number of comorbidities is another major risk factor, as 50% of the deceased patients were diagnosed with 3 or more comorbidities (Fig. 2B).

**Fig. 2.**
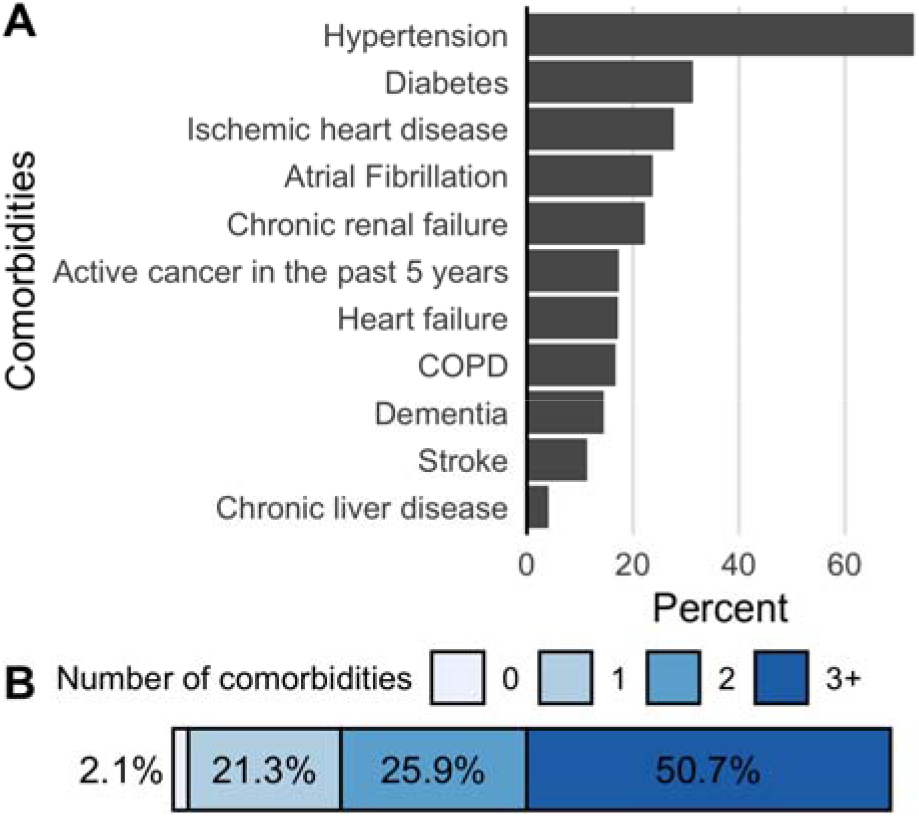
Comorbidities observed in deceased COVID-19 subjects. (A) Percentage of deceased patients diagnosed with the corresponding comorbidity. (B) Percentage of deceased patients with zero, one, two, and three or more comorbidities.

COVID-19 is not the only disease caused by pathogens and leading to severe and often deadly pneumonia. To get a broader view on the relationship between aging and lung disease, we analyzed the UK Biobank (UKB) dataset and observed that the incidence of pneumonia (the UKB disease code J18, “pneumonia, the organism is unspecified”) was also higher in men than in women and increased exponentially with age (Fig. 3A). The disease incidence doubling time was approximately 5 years for both sexes.

**Fig. 3.**
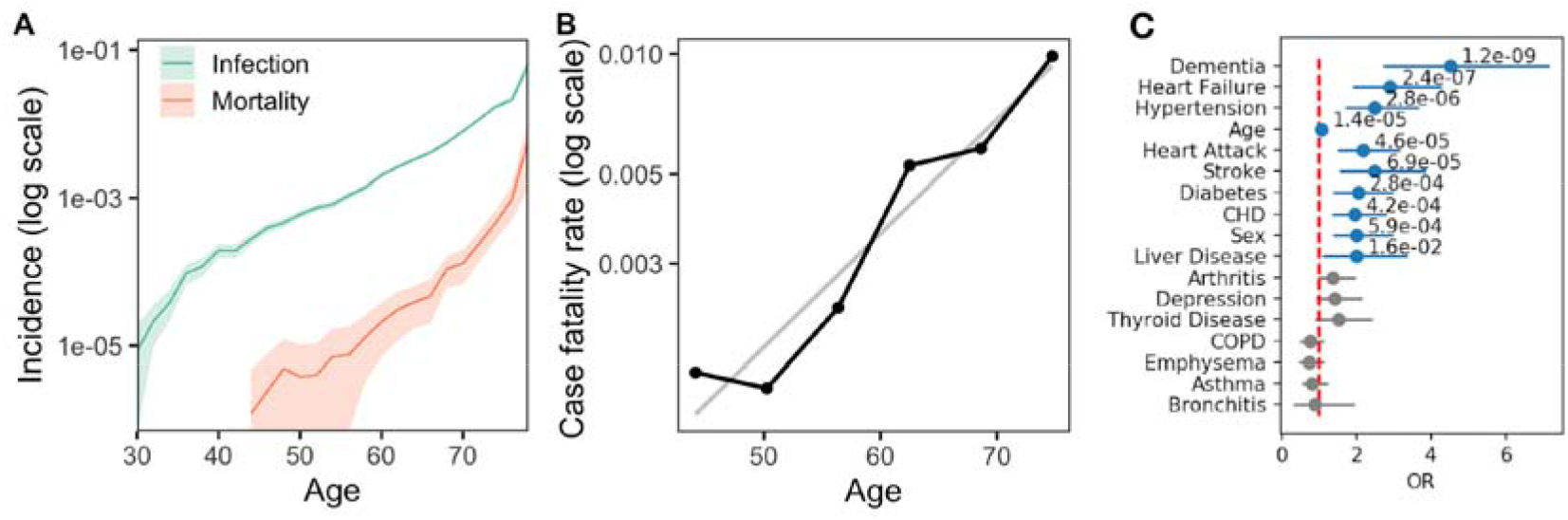
Pneumonia as an aging-related disease. (A) Incidence of pneumonia (green) and deaths with pneumonia as the primary cause (red) across age groups. (B) Case fatality rate as a function of age. Linear regression of log of case fatality rate on age is shown by a light line. (C) Risk factors associated with death from pneumonia. Logistic regression odds ratio (OR) for UKB phenotypes. Phenotypes with p-value<0.05 marked in blue. The data are from UKB and include both genders.

The disease cases covered under J18 label in the UKB were recorded by the hospitals. Overall, among 410,351 medical records, 10,109 participants had at least one episode of pneumonia recorded (18,493 in total), and in 104 patients pneumonia was listed as the primary cause of death. The pneumonia CFR grows exponentially for patients exceeding the age of 50 (Fig. 3B). The CFR doubling time was 9 years (with a large 95% CI from 5.6 to 12.4 years due to a low number of terminal cases), which is again compatible with both all-cause MRDT and COVID-19 CFR doubling time. The prevalence of age-related diseases was also a risk factor determining the probability of death from pneumonia. This could be seen from a series of univariate logistic models to determine the effects of age, sex, and prevalence of pre-existing conditions on the patient survival (Fig. 3C). The log-odds ratio (log-OR) associated with age was 0.056 (p<1e-5), which is equivalent to the CFR doubling time of approximately 10 years that is exactly the CFR doubling time in Fig. 3B. Thus, like COVID-19, pneumonia is a disease of aging.

It is not well understood why SARS-CoV-2 infection shows such a strong dependence on age. Several studies report on the role of ACE2 as the SARS-CoV and SARS-CoV-2 transmembrane receptor^16,20^. We hypothesized that age-dependent changes in the expression of ACE2 may contribute to the severity of the disease or to mortality of the elderly. A recent study analyzed ACE2 expression levels in various datasets with regard to confounding factors such as sex, smoking, age, and race^21^. Although it did not find an effect of aging, it is possible it may be due insufficient sample size. Therefore, we analyzed GTEx v8, the largest gene expression dataset of subjects across the adult lifespan, to assess the expression of the gene ACE2 across multiple tissues.

We found that the expression of ACE2 in the lung increases with age in subjects who were not on a ventilator at the time of death (Pearson’s r=0.23; p=0.0002; n=261) (Fig. 4A). This observation was consistent in both men and women (Supplementary Fig. 4). We further analyzed age-related changes in gene expression for all genes expressed in the lung. ACE2 exhibited one of the highest correlation coefficients with age in patients that were not on a ventilator (Fig. 4C). ACE2 shows increased expression in subjects on a ventilator not only in the lung but also in several other tissues (Supplementary Fig. 5). It is of interest that ACE2 is also upregulated by several drugs that are used to treat hypertension, a dominant pre-existing condition associated with COVID-19 mortality (Fig. 2). Furthermore, the expression of TLR7 in the lung is decreased with age, which may contribute to a poor immune response in the elderly (Fig. 4B). The expression of ACE2 as a function of age was analyzed in GTEx samples from subjects diagnosed with hypertension and other common COVID-19 comorbidities (Supplementary Fig. 6 and Supplementary Fig. 7). In addition, we evaluated the expression of genes related to the immune function in the lung and observed gender-associated differences, e.g. an age-related upregulation of pathways, such as innate immune response and negative regulation of adaptive immunity. Interestingly, the viral genome replication pathway was found to be upregulated (Supplementary Fig. 8).

**Fig. 4.**
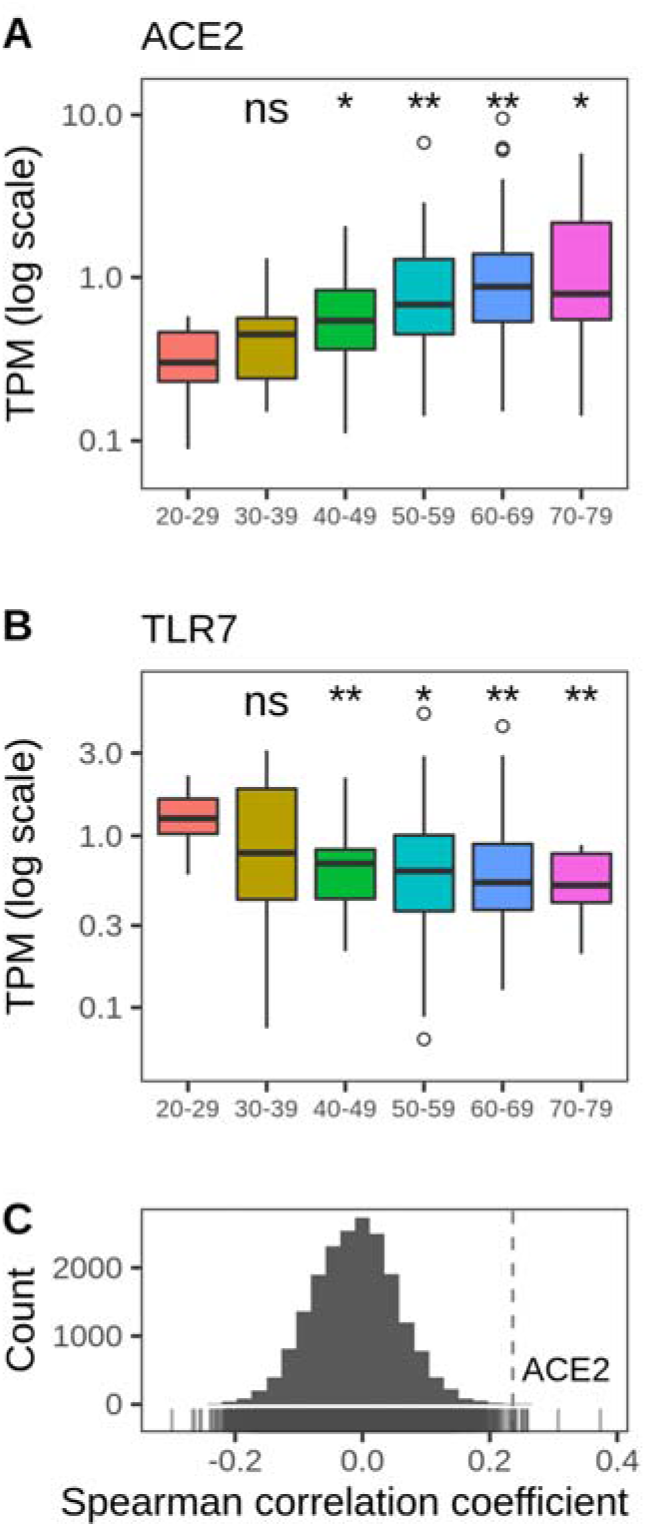
Age-related changes in SARS-CoV-2 receptor gene expression. Expression of (A) ACE2 (SARS-CoV-2 receptor) and (B) TLR7 in the human lung. P-value was calculated using Mann-Whitney test, ns - not significant, * p<0.05, ** p<0.01. (C) Distribution of genome-wide correlations between age and expression of genes in the lung. ACE2 is one of top genes whose expression increases with age. GTEx was used for these analyses. Cases on ventilator immediately before death were excluded.

While further analyses are needed, our study suggests that age-related changes in ACE2 expression at the entry point of the virus into the cells may contribute to the age-related increase in the SARS-CoV-2 infections and mortality.

## Discussion

Our study establishes COVID-19 as an emergent disease of aging. This conclusion is based on (i) an exponential growth of its CFR with age, (ii) the COVID-19 mortality rate doubling time approaching that of all-cause human mortality, (iii) higher mortality in men than in women, (iv) strong association with pre-existing age-related diseases, (v) COVID-19 being a subset of all-cause pneumonia, which is itself a disease of aging, and (vi) an age-related increase in the SARS-CoV-2 receptor mRNA expression in the lungs of non-ventilator subjects. Together, this puts COVID-19 in line with the archetypical diseases of aging, such as type II diabetes, cancer, Alzheimer’s disease, and cardiovascular diseases.

The SARS-CoV-2 infection rate also showed an age-related increase (Fig. 1A and Supplementary Fig. 2); however, this was primarily due to a low incidence among children and young adults, whereas its subsequent age-related changes were inconsistent. Moreover, testing priorities, age structure of the population and reporting procedures may bias this analysis (e.g., younger people may be asymptomatic and therefore tested less). Further studies are needed to assess the effect of age on the infection rate. Like incidence, CFR substantially varies among the countries (e.g., it is 10-fold higher in Italy and Spain than in Germany), which may also be explained by different strategies of reporting and testing. This, however, should not affect the age-related patterns in our study as we compare age groups within countries and not across them. Indeed, we observe similar COVID-19 mortality rate doubling times across all countries examined, even though they report drastically different incidences, testing and reporting strategies, and CFRs.

Age-adjusted all cause mortality is known to be higher in men than in women, and, as a result, women live longer than men. In the case of COVID-19, this trend seems to be even stronger. For example, across most age groups, men are twice more likely to die from COVID-19 than women in Italy (Fig. 1C), with the exception of one age group - 90 years old and older. However, the population age structure may explain the higher mortality in oldest women. Pre-existing conditions substantially increase the risk of death from COVID-19. These are primarily age-related diseases, i.e. hypertension, type II diabetes, and cardiovascular diseases. This risk is even higher in patients with multiple comorbidities. Overall, it further supports the notion that COVID-19 is an age-related disease.

COVID-19 patients die from severe pneumonia triggered by the specific virus infection. Many other pathogens are also known to cause pneumonia, which is particularly deadly for the elderly; however, older people are not always affected by viral infection or pneumonia, e.g. the largest pandemic in medical history - the Spanish influenza - affected young people (< 65 years old) disproportionately more severely, with the peak mortality at around 30 years (Supplementary Fig. 9). Based on UKB clinical histories, we investigated pneumonia as a proxy for all complications from respiratory diseases. The pneumonia incidence increased exponentially with age, so that the disease incidence doubled every 5 years. This is close to the all-cause MRDT from the Gompertz mortality law. Our observation of increasing incidence rate with age is consistent with previous studies of the incidence and MRDT of community acquired pneumonia in Germany^22^. The mortality associated with pneumonia increased faster in the elderly patients. The corresponding CFR increased exponentially with age for patients older than 50, and the CFR doubling time, again, was close to the Gompertz exponent. As in the case with COVID-19, risks of death from pneumonia were significantly higher in men and in patients with pre-existing conditions, such as hypertension, diabetes, and coronary heart disease, but not in patients with, e.g., COPD. A similar pattern was also seen in a recent study of the clinical course of COVID-19^23^. The common risk factors of severe pneumonia caused by COVID-19 and by infections in UKB suggest shared and pathogen-independent mechanisms. The appearance of the incidence and CFR acceleration rates close to the all-cause MRDT from the Gompertz mortality law is a signature of the first-order effects of aging. Together, this links COVID-19 and pneumonia as diseases of aging.

The association of COVID-19 CFR with aging as well as the association of the incidence and CFR of pneumonia with aging may open a way for using future anti-aging drugs, such as senolytics^24,25^, for both the prophylaxis and treatment of potentially deadly infectious diseases. First treatments seem to reduce markers of frailty and inflammation in pre- and clinical trials^26^ and hence may be helpful to prevent deadly complications. State-of-the-art biomarkers of aging such as DNA methylation based clocks^27^, could be used to stratify patients to define cohorts for expedite clinical trials and subsequently to select the most vulnerable individuals for treatment.

The mechanistic basis for the age-related pattern of COVID-19 mortality remains unclear. Our finding of an elevated age-related expression of ACE2 in the lungs of subjects with non-ventilator deaths may provide a clue. ACE2 is the site for the entry of SARS-CoV-2 into the cell. An age-related increase in the expression of this gene, together with the depletion of antiviral defenses, would naturally support a higher damaging effect of the coronavirus in the lung. It should be noted, however, that while ACE2 specifically promotes SARS-coronavirus infections, it also protects lungs from injury^28^. In addition, various tissues harbor different ACE2 gene expression levels and may account for complications other than pneumonia, such as diarrhea observed in a small sample of COVID-19 positive patients. At the protein level, lung and its alveolar type II cells were found to have low or undetectable ACE2 protein levels. Interestingly, ventilator cases showed no increase in ACE2 expression with age. The main difference between ventilator and non-ventilator cases is in the young subjects, wherein the expression of ACE2 in the ventilator cases is higher (Supplementary Fig. 10). The implications of variable expression patterns of ACE2 mRNA and protein across ages, tissues, and ventilator cases should be investigated in further studies.

Therapeutic strategies that support sequestration of the virus and protect lungs by a soluble form of ACE2 are currently under development^28^. These and other strategies should be particularly useful as the SARS-CoV-2 vaccine is not yet available. Moreover, vaccines are less efficient in the elderly, leading to a high rate of infections even in vaccinated individuals. For example, the yearly influenza vaccine is only 40% to 60% efficient in older individuals^29^. Therefore, targeting ACE2 may be viewed as both an immediate and a long-term strategy. However, as the case fatality rate grows with age, it should be possible to also adjust the pace of aging thereby targeting COVID-19. There are plenty of candidates for such a strategy^30^.

Overall, our analysis indicates that COVID-19 exhibits a clear characteristic of an age-related disease, and that age and age-related diseases are its major risk factors. Currently, there is a strong push towards therapeutic approaches targeting the machinery involved in viral biology. Our research suggests that targeting the aging process itself can be a viable orthogonal strategy against COVID-19 and other deadly respiratory diseases.

## Data Availability

This research has been conducted using GTEx data (dbGaP accession number phs000424.v8.p2) and the UK Biobank Resource under Application Number 21988

## Ethics approval

This research has been conducted using publicly available de-identified data: official reports on COVID-19 mortality listed in supplementary materials; GTEx data (dbGaP accession number phs000424.v8.p2); and the UK Biobank Resource under Application Number 21988. All relevant ethical guidelines had been followed.

## Competing interests

POF is a shareholder of Gero, and POF and AAZ are Gero employees.

## Acknowledgements

We thank members of the Gladyshev lab and the Gero team for discussion. Supported by NIH grants to VNG. This research has been conducted using the UK Biobank Resource under Application Number 21988.

## Notes

### Competing Interest Statement

POV is a shareholder of Gero, and POV and AAZ are Gero employees

